# A passive “OneHealth” surveillance system to track canine rabies in urban India

**DOI:** 10.1101/2022.02.23.22271029

**Authors:** Abi. Tamim Vanak, Neha Panchamia, Indrakshi Banerji, Malvika Nair

## Abstract

Robust and widely implementable “OneHealth” surveillance systems are required to detect and control the spread of zoonotic infectious diseases. Using technological tools coupled with a rapid field response and rapid diagnostics can help in surveillance of deadly diseases such as rabies, even in densely populated urban areas. Here, we describe the use of an animal rescue system (Hawk Data Pro) that was adapted as a passive surveillance tool for rabies in a large metropolis in India. We used a webline and helpline that reported injured or sick animals to an animal rescue facility to determine possible rabies cases in street animals in Pune city in western India. Suspected rabid animals were tested using lateral flow assays and this information was used to direct awareness materials on rabies as well as in conducting mass dog vaccinations in areas that reported multiple cases. Over a ∼4 year period, we received over 90,700 calls or reports, of which 1162 calls were for suspected rabies cases in dogs, and 6 for other animals (cats, goats and cattle). Of these, 749 dogs and 4 other animals tested positive for rabies. Most of the cases were reported from the densely human populated central Pune region. In response, ∼21,000 people were provided with educational materials on post-bite management, and ∼23,000 dogs were vaccinated. We show that the adoption of the Hawk Data Pro system as a passive surveillance tool allowed us to document an ongoing outbreak of rabies in a large metropolis in India. Such systems can be modified or adapted to other areas as well to meet the surveillance and reporting requirements of the WHO’s Zeroby30 strategy to eliminate rabies globally.

## Introduction

There is an increasing global demand for the establishment of responsive and scientifically sound surveillance systems to better understand and possibly predict outbreaks and spread of zoonotic infectious diseases (Halliday et al., 2017; Kelly et al., 2017; Morse et al., 2012). Surveillance systems need to be strengthened, particularly in developing countries (Halliday et al., 2017; WHO 2012), so that the emergence and spread of zoonotic pathogens can potentially be decelerated or prevented with timely detection and disease reporting (Halliday et al., 2012; Loh et al., 2015; Vrbova et al., 2010). Unfortunately, most efforts towards strengthening response to zoonoses have been focused mainly on technical and diagnostic capacity building and training, with very little emphasis on establishment of robust systems for field data collection (Halliday et al., 2011).

The case of rabies in India is a classic example of such neglect in surveillance mechanisms (Kakkar et al., 2012). Rabies is still considered a neglected disease (WHO/WSPA 1990) in India despite the country having more cases of human rabies deaths than any other in the world (Hampson et al., 2015; Knobel et al., 2005). Even today, the primary citation for rabies incidences in India comes from studies carried out in 2003 (Sudarshan et al., 2007). India accounts for approximately 36% of human rabies cases worldwide and is a major focus for global efforts to eradicate rabies by 2030 (WHO 2021). More than 95% of rabies cases are caused by free-ranging domestic dogs (FRD). However, surveillance of rabies in its primary reservoir host is limited, both due to a lack of field capacity as well as diagnostic facilities (Taylor et al., 2017), a common problem in most developing countries (Wallace et al., 2017). One of the key reasons for this grim reality is the lack of a comprehensive policy for rabies control that is based on rigorous scientific study of disease dynamics in India (NCDC 2020).

The Global Alliance for Rabies Control’s Blueprint for Rabies Elimination recognises the importance of robust surveillance and diagnostic capabilities to better direct post-bite management resources (OIE 2018). Such surveillance would also allow for evaluation of interventions such as mass anti-rabies vaccinations, or the impact of increased awareness to the human population. Thus far in India, passive surveillance has been limited to ad-hoc reporting of suspected animals to veterinary diagnostic facilities, usually in urban centres. This has resulted in only broad-scale patterns of rabies incidences in dogs and other animals becoming apparent, and high rates of under-reporting of cases (Brookes et al., 2016; Gill et al., 2019).

Here we report the use of a novel street animal rescue management system which was adapted to function as a systematic passive “One Health” surveillance of rabies in a large Indian metropolis. We demonstrate how this system was able to detect a large-scale ongoing rabies outbreak in the city through reporting, response and testing using rapid diagnostic kits. This system was able to direct concerted awareness efforts for both post-exposure prophylaxis for human patients and conduct mass-vaccination for FRD. Furthermore, through this system, we were able to highlight the scale of rabies prevalence in one of the largest cities in India, and the challenges of achieving Zeroby2030.

## Method

### Study Area

Pune city, located in the state of Maharashtra in western India is a large metropolis that has a population of 7.4 million (India Population 2020). There have been no systematic surveys of the dog population in this city, but it is estimated that the dog population is ∼3,00,000 (Hindustan Times 2020). Several anti-rabies vaccination drives of varying scales have been conducted from 2010 to 2020 in the city by non-governmental organizations and local groups, however, there is inadequate information on the locations or whether the same animals were provided annual booster shots. An animal rescue organization called RESQ Charitable Trust (RESQCT) which had been operating in the city since 2007, began a Rabies Response Unit in 2017 which systematically responded to rabies suspected cases, dedicated a quarantine and testing facility as well as conducted mass anti-rabies vaccination drives (RESQCT).

### Reporting Systems

RESQCT receives reports of animal emergencies and potentially rabid animals through two contact points – one, telephonically through their ‘Helpline’ and two, online through their ‘Webline’. The RESQCT helpline number is well distributed across the city for over a decade through several media, online and offline, and is the primary number provided by the Dog Control Cell of the city’s municipal corporation in response to any complaint regarding aggressive or biting dogs. RESQCT has a ‘Report’ button placed prominently on their website and since 2016, several awareness efforts, online and offline, have been engaged to popularize the use of this service to the general public to report any hurt, injured or sick animal.

#### Helpline Reporting

The RESQCT Helpline is a mobile number that is kept operational from 10:00 am to 00:00 midnight i.e., 14 hours per day, 365 days a year and is handled by personnel trained in responding with a detailed situation- and symptom-based questionnaire to identify suspected rabies cases.

##### Webline Reporting

The animals reported on the RESQCT Webline are systematically listed on the Webline section of the ‘Hawk Data Pro’ (<u>QiCode Solutions)</u> which is a web-based online management system for Animal Rescue Organizations that enables citizen reporting, transport and task assignment, animal data management, team and donor management and generates custom reports (Fig 1).

**Fig. 1.**
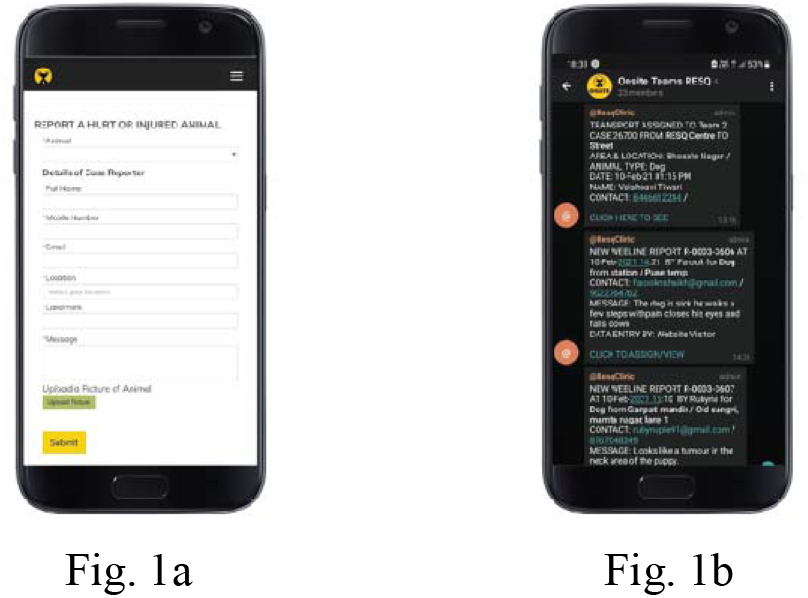
When a citizen wants to report an injured or sick animal, the RESQCT website has a ‘Report a Case’ prominently positioned on the top. **Fig. 1a**. Shows the screen and fields filled in by the citizen reporting the animal. **Fig 1b**. Every submission triggers a message to the RESQCT team mobile application (Telegram App via an API).

The report or entry is analyzed by trained personnel and based on the information submitted, a telephonic call is made to the reporter to request more details and plan further response.

### Response

Based on the information provided by the reporter, the Rabies Response Unit Ambulance is immediately sent out to capture the suspected animal. In doubtful cases, an additional video is requested to be sent for verification. A rabies suspected dog is captured using a butterfly net and placed inside a fiber-crate for transport to the RESQ Rabies Quarantine Unit. All necessary PPE is used during capture and all equipment used to handle animals is disinfected immediately after the transfer is complete.

If the report indicates that the animal has bitten an individual (or more), the veterinarian in charge of the Rabies Unit telephonically conveys the importance of taking post-exposure prophylaxis measures to any and all affected individuals. If any other street dogs in the vicinity have been bitten by the suspected animal, the RESQCT team vaccinates the free-ranging dogs (if present) in and around the capture location, distributes rabies awareness materials, and alerts local citizens to be vigilant and report any changes in behavior of the free-roaming dogs present in the area.

### Quarantine and Testing

Animals exhibiting two or more symptoms of rabies, or a history of aggression/ bites were brought into the rabies ward and isolated on admission. Symptoms include hyperesthesia, hyper-aggression (towards humans, other animals or objects), paralysis of mandible, paralysis of larynx, inability to swallow, and other signs associated with rabies.

Suspected rabid animals were tested using commercially available rapid antigen testing kits. An ante-mortem saliva swab sample was collected and a post-mortem brain sample was collected by inserting a thick plastic straw through a small insertion at the base of the neck into the foramen magnum, towards the frontal bone (Yale et al., 2019). The samples were then processed following manufacturer’s instructions provided for the Bionote Rapid Rabies Antigen Test Kit (BIONOTE). The Bionote antigen lateral flow assay (LFA) kits have been found to perform well and are generally recommended for rapid field tests (Yale et al., 2019; Servat et al., 2019), though with the caveat that they need to be cross validated (Eggerbauer et al., 2016; Klein et al., 2020). A portion of each sample was frozen at -20°C and then transported on ice for confirmatory marker-gene based analysis using reverse transcriptase polymerase chain reaction at either the WHO accredited laboratory at the National Institute of Mental Health and Neuro-Sciences (NIMHANS), Bangalore or to the Indian Institute of Science, Bangalore (Vanak, A. T. et al., forthcoming).

### Data Management

Any notable behavioral data was recorded once the animal was reported, until death. Information was entered in the Hawk Data Pro system and a symptom log along with photographs or videos was maintained independently for each animal as well.

### Local Community Awareness Outreach and Action post rabies positive testing

In case of an animal that had tested positive for rabies with the LFA test, the local community was made aware and provided with further information on post-exposure prophylaxis and other safety measures. The location information was recorded and used for geographic mapping of the positive rabies case to determine ‘Rabies Outbreak Zones/Hotspots’ in the city. Mass anti-rabies vaccination drives were conducted in these areas that were identified as hotspots of rabies.

### Analysis

We generated descriptive statistics of the reported suspected and positive cases, and summarised them by gender and month.

## Results

During September 2017 to July 2021, the RESQCT helpline and weblines received over 90,700 calls or reports, of which 1162 calls were processed as suspected rabies cases in dogs (Males = 585, females = 553, unknown = 24), cats (n= 2), caprine (n = 1) and bovines (n = 3).

Out of these, 749 dogs (males = 360, females = 377, unknown = 12) tested positive for rabies with the LFA kit. Proportionally, more females (68.2% of 553) than males (61.5% of 585) tested positive amongst those reported (Fig. 2). 121 suspected rabid dogs were neutered (10.5%), of which 80 tested positive. Furthermore, 4 dogs with a history of recent vaccination (i.e. dogs that were known to be vaccinated by either RESQ or a guardian within the last 6-12 months), were reported, of which 100% were positive for rabies. Forty pet dogs were reported, of which 38 tested positive for rabies. Of the 3 suspected bovine and 1 caprine case, all tested positive and one of the two cats tested positive.

**Figure 2.**
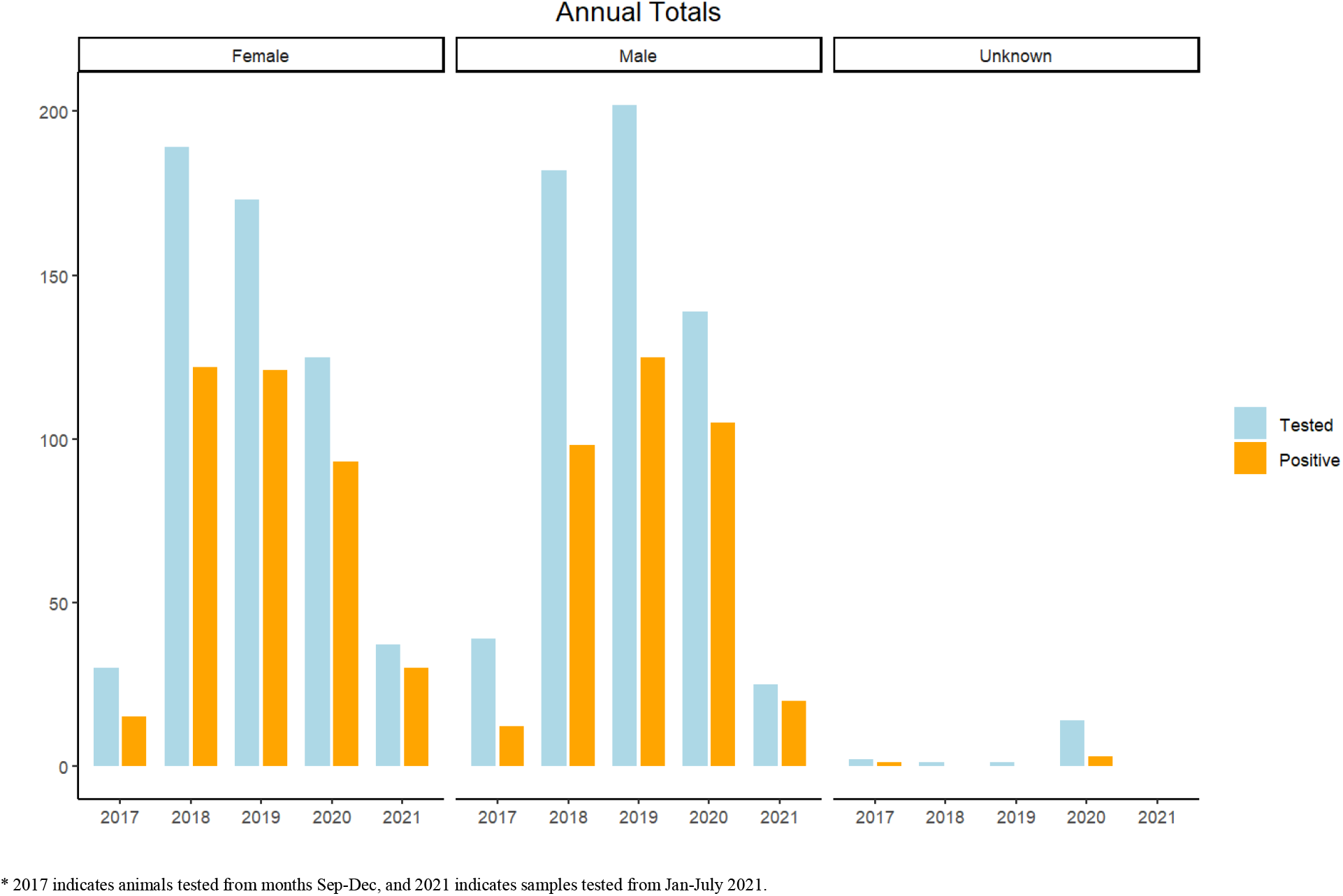
Total animals suspected of rabies and tested positive during this study. (Sep 2017-Jul 2021). Overall, more male animals were reported, but proportionally more females tested positive.

Nearly 4% of all dogs that tested positive for rabies came with a reported history of biting either people or other dogs. In one case, a rabid dog had bitten seven people over the course of two days (Hindustan Times 2017). The number of rabid cases were similar between the years 2018-2020 (Fig 2). Rabies cases peaked between December to January and were lower in the summer months (Fig. 3). The highest number of rabies cases year on year were reported from the high human density old town region of central Pune (Fig. 4).

**Figure 3.**
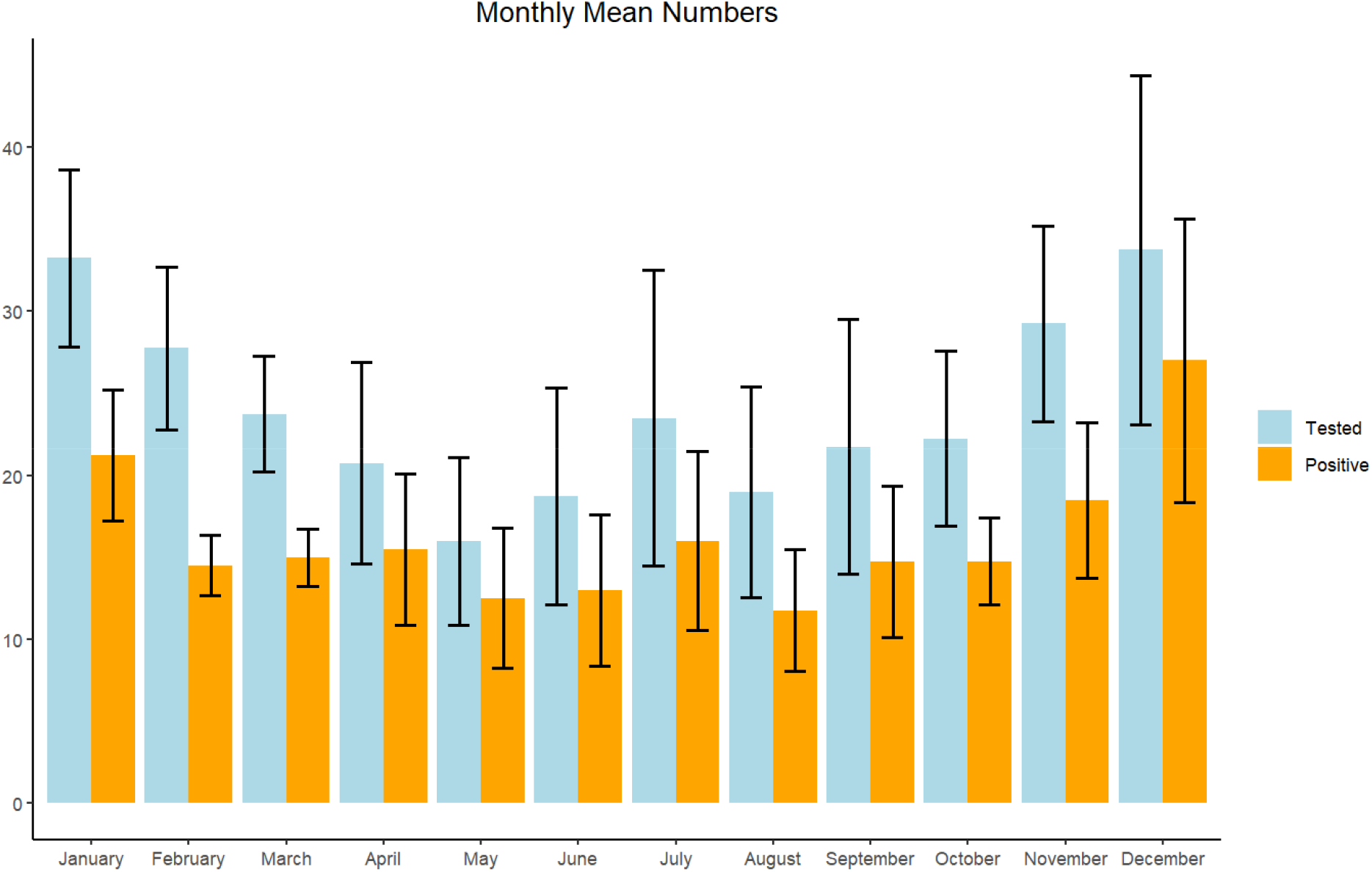
The mean monthly number (+-SE) of rabies cases in Pune city shows a seasonal trend with the cooler months of January and December showing a marginal trend of higher number of cases compared to the rest of the year.

**Figure 4.**
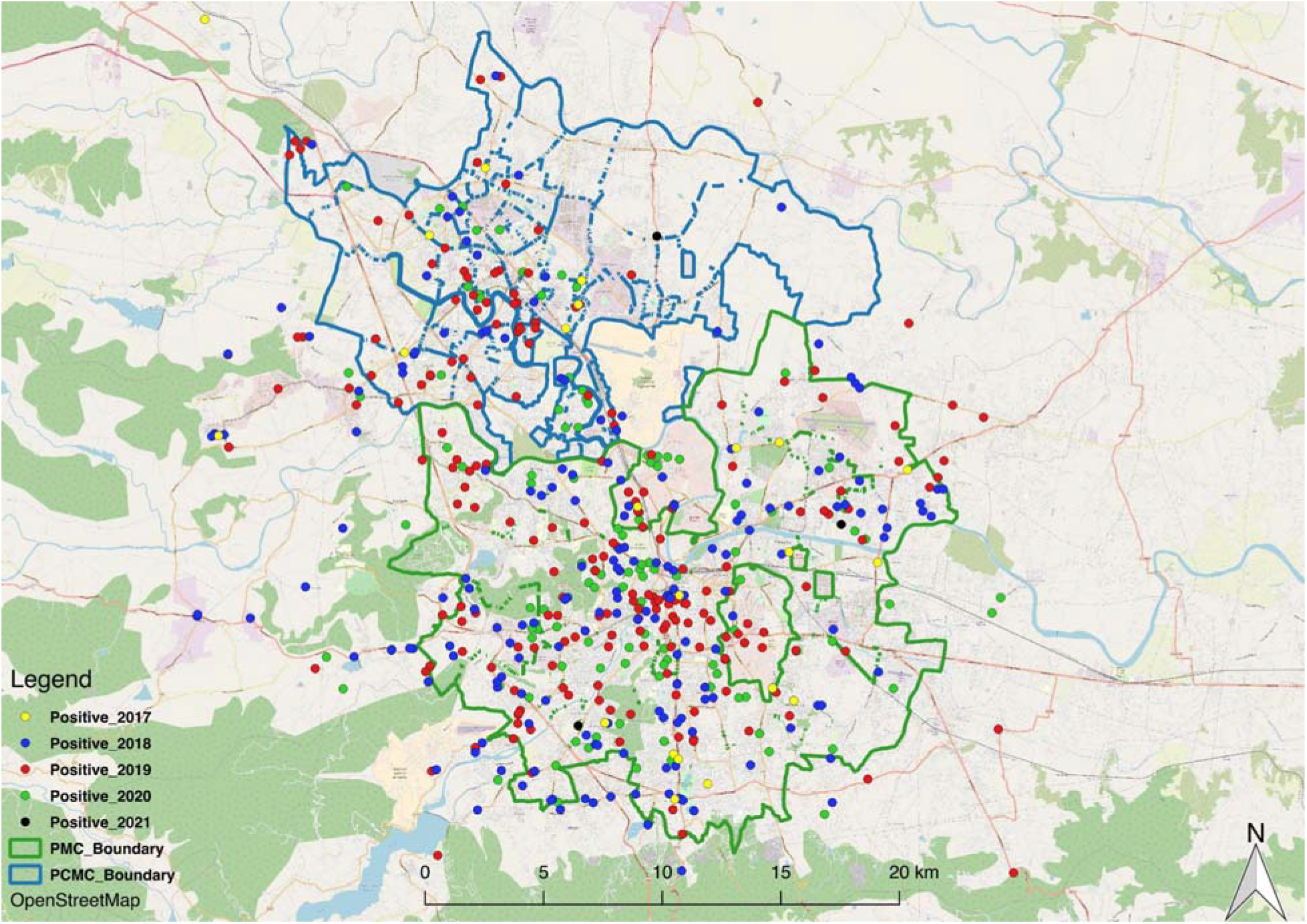
Distribution of detected rabies cases in Pune city and adjoining peri-urban areas during the study period. Most cases were detected in the old city area of central Pune, as well as in central Pimpri-Chinchwad Municipality limits to the NorthWest.

As a follow-up action to suspected rabid dogs being reported from an area, 20,730 people were provided with awareness materials and advised to seek PEP immediately if they were exposed. Targeted and mass ARV campaigns were conducted and ARV was administered to 22,804 dogs from 2018 to October 2021.

## Discussion

The WHO’s ‘Zero by 30’ goal to eliminate rabies worldwide stresses on the importance of generating data to measure the success and impact of control and surveillance strategies, and to adapt them for local use. While the method described in this paper was essentially a passive surveillance method, it resulted in the creation of a system that encompasses four main steps - ‘reporting’, ‘responding’, ‘testing’ and conducting ‘outreach’. First, through clinical presentation and diagnostic tests, we accounted for potential asymptomatic or cross-symptomatic cases, which, when ignored, lead to underreporting of the disease. We then conducted vaccination and awareness drives in the localities with a high canine rabies incidence rate, potentially reducing human cases.

Adapting the RESQCT ‘reporting’ system Hawk Data Pro database as a passive surveillance tool in Pune city allowed us to capture the extent and severity of an ongoing canine rabies outbreak in a densely populated Indian city. These cases represent amongst the highest number recorded in any Indian city consistently over several years, and helps to highlight the scale of the canine rabies problem in India. Outbreaks of similar magnitude have been reported from major urban areas in other parts of the world over two decades ago (e.g. see Bolivia and Mexico (Eng et al., 1993; Widdowson et al., 2002)), indicating that efforts to control canine rabies in India still have a long way to go.

A majority of the dogs that were reported and tested positive for rabies were free-ranging dogs that were presumably unowned, partially owned, or so-called “community-owned”. This suggests that although there have been ongoing efforts at controlling rabies in Pune city (https://timesofindia.indiatimes.com/city/pune/civic-body-steps-up-to-check-stray-dog-population/articleshow/86591746.cms), this has largely been in-effective (Belsare and Vanak, 2020). The only government mandated program to control rabies is the Animal Birth Control program (a form of CNVR) which administers anti-rabies vaccination to a dog captured for sterilization or neutering. Such dogs are typically marked with an ear-notch. There is no provision of periodic revaccination or booster shots for dogs that have been sterilised. This lack of an effective re-vaccination program is evident, as nearly 11% of dogs that tested positive were neutered and thus had received at least one shot of ARV. Furthermore, 4 dogs that had a recent history of vaccination (<1 year) also tested positive, indicating either that they were already infected at the time of vaccination, case of importation of latent dogs or failed to sero-convert (Laager et al., 2019; Van Heerden et al., 2002). Worryingly, 5% of dogs that tested positive were owned pet dogs (many of which were pedigrees) presumably under some form of confinement. This suggests that there is a general lack of awareness about rabies, even amongst urban pet owners about the need for regular vaccination. This is similar to the findings of Tiwari et al. (2019), who reported an alarmingly high proportion of urban dog owners (∼30%) who did not vaccinate their dogs.

There seemed to be a seasonal trend in reported rabies cases, with a peak post-monsoon in the winter months. Several studies have shown peaks in breeding activity of dogs during the monsoon season, with high rates of congregation and aggression between animals (Chawla and Reece, 2002; Fielding et al., 2021; Pal, 2001). Incidences of rabies have also been correlated with this seasonal pattern in breeding, potentially due to greater movement of dogs and higher contact rates (Brookes et al., 2016). The time-series information in the annual number of rabies cases was not sufficient for us to deduce any generalised pattern of increasing, decreasing or stable trends. It is possible that the reduced movement of people on the streets due to the Covid19 pandemic induced lockdowns may have resulted in fewer dogs being reported in 2020 and early 2021. However, a longer-term monitoring is required to better understand these temporal patterns.

The percentage of suspected animals that tested positive also indicates that besides surveillance and providing a means for ‘reporting’ information about suspected animals, a crucial step in the management of rabies on ground i.e., ‘responding’ greatly depends on analysis of reports received and rapid response. In the case of Pune city, RESQCT’s trained dog catching team as well as a quarantine and testing facility enabled the management of suspected rabies cases in the city. Furthermore, it also allowed a rapid deployment of awareness materials to the “at-risk” human population, and targeted vaccination campaigns for dogs that were potentially exposed to rabies. Since one of the primary causes of human rabies deaths is a lack of information about the proper procedure after a bite (Masthi et al., 2019; Tiwari et al., 2019), including administration and dosage rates of the PEP, awareness plays a big role in controlling this disease. Although it is difficult to establish direct causation, there were no reports of rabies in humans that originated from exposure within Pune city for the years that this program was implemented. Similarly, Bangladesh, since 2011, has shifted the focus to dog-bite management and mass dog vaccination, which has reduced the human-rabies deaths significantly (Hossain et al., 2020). Surveillance data can help estimate the impact of disruption of vaccination drives, possible animal infections and human rabies exposures, and costs for human post-exposure prophylaxis (Kunkel et al., 2021). A real-time surveillance program and response system are therefore the first steps in the Stepwise Approach to Planning and Evaluation in the Canine Rabies Blueprint (https://caninerabiesblueprint.org/ accessed on 10/10/20).

A National Action Plan for dog-mediated Rabies Elimination (NAPRE) in India by 2030 has been introduced by the Ministry of Health & Family Welfare (NCDC) in alliance with the Ministry of Fisheries, Animal Husbandry, and Dairying, Government of India in September 2021 (NAPRE 2021). The program aims at reducing human deaths due to canine rabies by undertaking mass dog vaccinations and providing timely post-exposure treatment. The program relies on the missing inter-sectoral action and hence there is hope that the fragmented and uncoordinated activities by various stakeholders will align effectively with the newly proposed One Health approach for elimination of rabies in this roadmap (OIE 2020). The systematic passive surveillance system described in this study can be adopted in the One Health surveillance of rabies across most cities and rural areas because the web-based reporting system, Hawk Data Pro, requires only a basic browser and limited internet connectivity. In the absence of a response team to capture a suspected animal or having quarantine and testing facilities available, there is still great value in implementing a surveillance system to determine thresholds of potential concern and develop adaptive management frameworks to aid governmental bodies to initiate action and allocate resources (training and infrastructure for rabies control and management) specifically into regions that require attention.

We recognise that this is a method of passive surveillance and relies on data collected ‘per-chance’. Although it allows for surveillance of rabies in large urban settings, this method may still under-report actual incidence rates. However, combining this method with more robust active surveillance can provide better estimates of the burden of rabies in street dog populations in urban areas.

## Conclusion

Strengthening surveillance systems is integral to address the underreporting of rabies, and to achieve the target of eliminating rabies by 2030 in India. Data generated through such effective surveillance can help inform about rabies by following the standard case definitions of suspected, probable and confirmed cases as described under the National Rabies Control Program and incorporated at the Integrated Disease Surveillance Portal that is working to make rabies a notifiable disease (NCDC 2022). The passive surveillance approach reported here allowed us to identify potential hotspots of rabies and ensured timely provision of awareness materials and PEP to the identified at-risk population. Helpline and Web portal-based notification of animal rabies can serve both human and animal health components. The approach can be adapted for continuous OneHealth rabies surveillance and is well aligned with the newly designed objectives of NAPRE, a plan developed with consideration for intersectoral planning, operational research and based on the One Health approach (NAPRE 2021).

## Data Availability

All data produced in the present study are available upon reasonable request to the authors

## Acknowledgement

We thank the veterinarians, staff and volunteers of ResQ CT, (Pune, India) for their active participation in this study. We thank N. B. Jadeja, A. Katna and A. Kulkarni for help with preparing the manuscript. This study was funded through a DBT/Wellcome Trust India Alliance grant to ATV (Grant number: IA/CPHI/15/1/502028).

## Ethics statement

The authors confirm that the ethical policies of the journal, as noted on the journal’s author guidelines page, have been adhered to and this study has been approved by the ATREE Animal Ethics Committee (AAEC/101/2016). All animals that were housed at the ResQ CT hospital follow the guidelines set by the Animal Welfare Board of India.

